# Development and Internal Validation of Models Predicting the Health Insurance Status of Participants in the German National Cohort

**DOI:** 10.1101/2024.04.09.24305544

**Authors:** Ilona Hrudey, Enno Swart, Hansjörg Baurecht, Heiko Becher, Antje Damms-Machado, Wolfgang Hoffmann, Karl-Heinz Jöckel, Nadja Kartschmit, Verena Katzke, Thomas Keil, Bianca Kollhorst, Michael Leitzmann, Claudia Meinke-Franze, Karin B. Michels, Rafael Mikolajczyk, Tobias Niedermaier, Iris Pigeot, Sabine Schipf, Börge Schmidt, Barbara Walter, Stefan Willich, Robert Wolff, Christoph Stallmann

**Affiliations:** Institute of Social Medicine and Health Systems Research, Otto-von-Guericke University Magdeburg, Faculty of Medicine, Magdeburg, Germany; Institute of Epidemiology and Preventive Medicine, University of Regensburg, Faculty of Medicine, Regensburg, Germany; Heidelberg Institute of Global Health, University Hospital Heidelberg, Heidelberg, Germany; Max Rubner-Institut (MRI), Bundesforschungsinstitut für Ernährung und Lebensmittel, Institut für Kinderernährung, Karlsruhe, Germany; Institute for Community Medicine, University Medicine Greifswald, Greifswald, Germany; Institute of Medical Informatics, Biometry and Epidemiology, University Hospital of Essen, Essen, Germany; Institute for Outcomes Research, Center for Medical Data Science, Medical University of Vienna, Vienna, Austria; German Cancer Research Centre DKFZ, Heidelberg, Germany; Institute for Social Medicine, Epidemiology und Health Economics, Charité - Universitätsmedizin Berlin, Berlin, Germany; Institute for Clinical Epidemiology and Biometry, University of Wuerzburg, Wuerzburg, Germany; State Institute of Health, Bavarian Health and Food Safety Authority, Bad Kissingen, Germany; Leibniz Institute for Prevention Research and Epidemiology - BIPS, Bremen, Germany; Institute for Prevention and Cancer Epidemiology, University of Freiburg, Faculty of Medicine and Medical Centre, Freiburg, Germany; Institute of Medical Epidemiology, Biometrics and Informatics, Martin-Luther-University Halle-Wittenberg, Halle, Germany; University of Bremen, Faculty of Mathematics and Computer Science, Bremen, Germany; Cancer Registry Saarland, Ministry of Labour, Social Affairs, Women and Health, Saarland, Germany

**Keywords:** prediction models, missing values, health insurance status, cohort study, primary data

## Abstract

**Background:** In Germany, all citizens must purchase health insurance, in either statutory (SHI) or private health insurance (PHI). Because of the division into SHI and PHI, person insurance’s status is an important variable for studies in the context of public health research. In the German National Cohort (NAKO), the variable on self-reported health insurance status of the participants has a high proportion of missing values (55.4%). The aim of our study was to develop and internally validate models to predict the health insurance status of NAKO baseline survey participants in order to replace missing values. In this respect, our research interest was focused on the question to which extent socio-demographic characteristics are suitable for predicting health insurance status.

**Methods:** We developed two prediction models including 53,796 participants to estimate the probability that a participant is either member of a SHI (model 1) or PHI (model 2). We identified eight predictors by literature research: occupation, income, education, sex, age, employment status, residential area, and marital status. The predictive performance was determined in the internal validation considering discrimination and calibration. Discrimination was assessed based on the Area Under the Curve (AUC) and the Receiver Operating Characteristic (ROC) curve and calibration was assessed based on the calibration slope and calibration plot.

**Results:** In model 1, the AUC was 0.91 (95% CI: 0.91-0.92) and the calibration slope was 0.97 (95% CI: 0.97-0.97). Model 2 had an AUC of 0.91 (95% CI: 0.90-0.91) and a calibration slope of 0.97 (95% CI: 0.97-0.97). Based on the calculated performance parameters both models turned out to show an almost ideal discrimination and calibration. Employment status and household income and to a lesser extent educational level, age, sex, marital status, and residential area are suitable for predicting health insurance status.

**Conclusions:** Socio-demographic characteristics especially employment status and household income assessed at NAKO’s baseline were suitable for predicting the statutory and private health insurance status. However, before applying the prediction models in other studies, an external validation in population-based studies is recommended.

## Introduction

With 205,264 participants, the German National Cohort (NAKO; German: *NAKO Gesundheitsstudie*) is the largest German population-based prospective cohort study to date. The primary goal of the NAKO is to investigate the aetiology, risk, and protective factors of widespread chronic and infectious diseases such as cancer, diabetes mellitus, neurodegenerative and psychiatric diseases as well as diseases of the cardiovascular and respiratory systems. The findings will be used to derive new strategies for the prevention, early detection, and treatment of these diseases. In addition to the elicitation and collection of comprehensive health data, a sustainable infrastructure for public health research will be established in Germany by this huge cohort [1–5]. As part of the passive follow-up, the collected primary data are enriched with claims and registry data (e.g. health insurance, pension insurance, and cancer registry data), which include information on the exposure and disease status as well as on the utilisation of medical services of the study participants [4, 6]. For the first time in Germany, record linkage of data from statutory (SHI) and private health insurances (PHI) with primary data of study participants will be realized [2, 6, 7].

In Germany, health insurance has been mandatory since 2009, i.e. all citizens must insure themselves either in the SHI or in the PHI. Cover through SHI is mandatory for employees and other groups (e.g. pensioners) with a gross income below the opt-out threshold (64,350€ per year in 2021). Persons with an income above the threshold can purchase substitutive PHI. Self-employed can choose between voluntary membership in the SHI and substitutive coverage through PHI, regardless of income. For certain professional groups (e.g. civil servants), membership in PHI is mandatory. In Germany, about 85% of the population are covered by SHI and 11% are covered by substitutive PHI. Sector-specific governmental schemes provide coverage for certain population groups such as police officers, soldiers and refugees. The coexistence of SHI and PHI leads to inequalities due to differences in financing, access and provision of health care [8, 9]. More details on the German health insurance system can be found in [8, 9].

Various studies have shown that health status, medical care and the distribution of socio-demographic characteristics differ between people with SHI and PHI. For example, privately insured people earn a higher average income and are on average healthier than statutory insured [7, 10–17]. Therefore, person insurance’s status is an important variable for studies in the context of public health research in Germany. However, existing studies that investigated differences between statutorily and privately insured persons are mainly cross-sectional and were subject to limitations such as small sample sizes in which subgroup analyses are difficult [18]. Also, claims data analyses have mostly used data from SHI [7]. Thus, approximately 11% privately insured persons of the German population [9] were ignored in most analyses [7]. In this respect, the NAKO offers a unique opportunity since health-related factors of statutorily and privately insured persons can be longitudinally analysed in a huge study population including a large number of collected variables [18–20].

## Background

The acquisition and scientific use of claims and registry data and its individual linkage with primary data in the NAKO requires informed consent, which is retroand prospectively valid for 5 years and must then be renewed [21]. Health insurance number, name of the insurance company and the information ’privately insured’ (yes/no) were recorded during the consent process [22, 23] from those participants who gave their informed consent (n=188,974; 92%; **Fig. 1**).

**Fig. 1.**
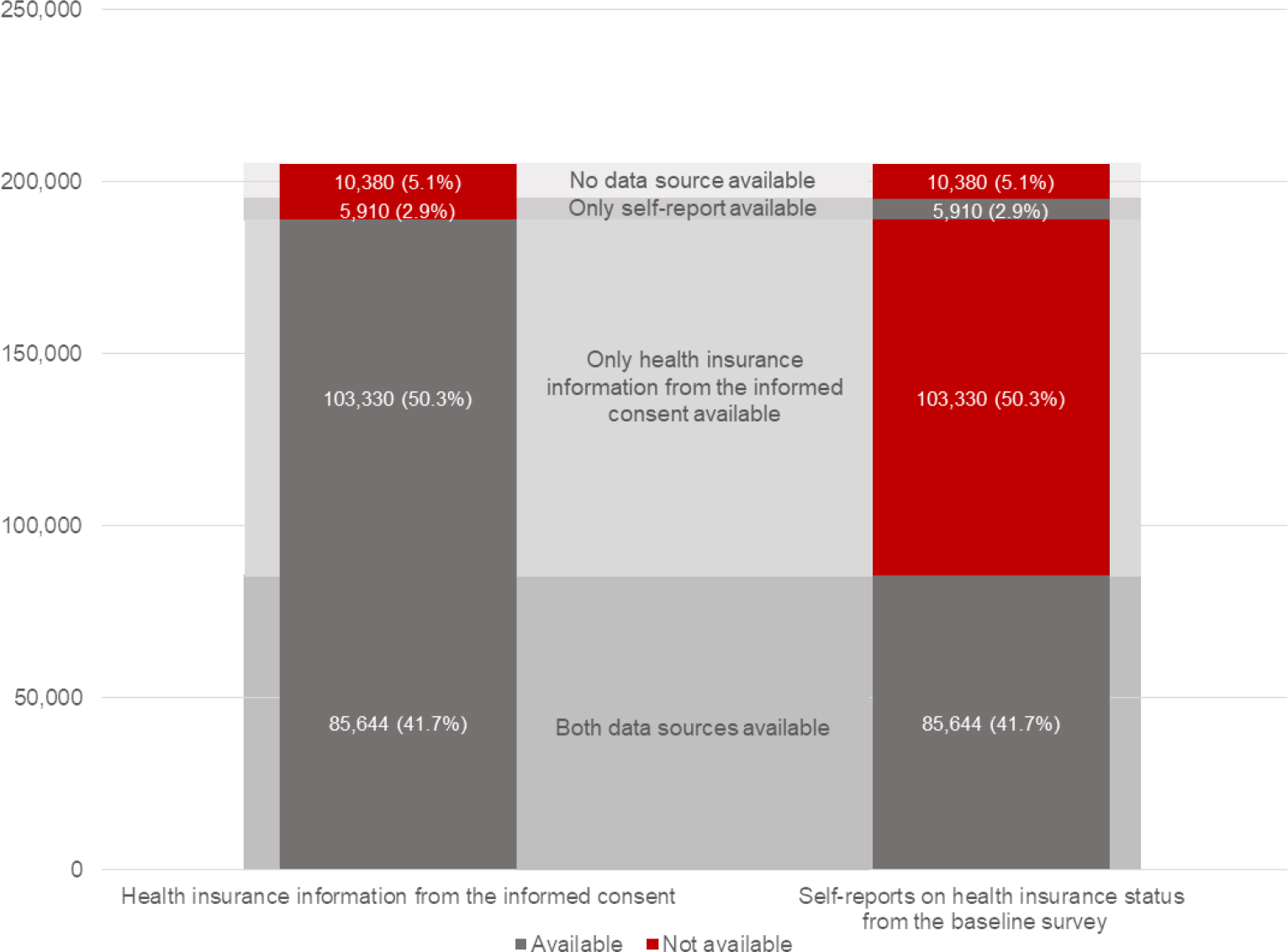
Completeness of data on health insurance status in the NAKO baseline assessment

To enable the comparison of health-relevant aspects between participants with SHI or PHI without actually having access to their claims data, the health insurance status has been additionally recorded since 2017 in the baseline survey on participants’ self-report. **Table 1** illustrates the recording of health insurance information in the NAKO. The baseline survey began in 2014. The question on health insurance status was subsequently included in 2017 as part of the revision of the touchscreen self-filler questionnaire. This resulted in the high number of missing values in the variable (n=113,710; 55.4%). For 10,380 participants (5.1%), no information on health insurance status is available in either data source (**Fig. 1**). In the NAKO’s follow-up (2018-2023), the health insurance status of all participants will be continuously recorded in a computer-assisted personal interview (CAPI).

**Table 1.** Recording of health insurance information in the NAKO baseline assessment (information on consent process from. [23])

| Consent process |  |
| --- | --- |
| Question | Answer option |
| Scanning of the health insurance card and determining the following information: |  |
| 'Privately insured' | Yes<br>No |
| Health insurance number | Free text |
| Health insurance number not brought along | Field to tick off |
| Number of the health insurance company | Free text |
| Name of the health insurance company | Drop-down list |
| Remarks on health insurance | Free text |
| Touchscreen self-filler questionnaire |  |
| Question | Answer option |
| Are you a member of a health insurance?* | Yes, I am a member of a SHI<br>Yes, I am a member of a PHI<br>Yes, I am otherwise insured<br>No, I am not insured<br>I don't know<br>Not specified |

\* Translation by authors

For the analysis of health-relevant differences between statutorily and privately health insured persons using the data set of the NAKO baseline survey, valid and non-missing information on the health insurance status is required. Incorrect information may result from the participants’ limited institutional knowledge of the German health insurance system. For example, it is conceivable that respondents claim to be a member of PHI although they have a supplementary PHI or are insured through sector-specific governmental schemes such as the *Freie Heilfürsorge*, which e.g. covers soldiers and police officers [24]. Using the incorrect self-report of health insurance status in a statistical analysis may introduce information bias by measurement error and by this may lead to biased estimators and, therefore, invalid study results [25, 26].

Self-reported health insurance status was already validated as part of the quality assurance of the baseline survey. For this purpose, the self-reported health insurance status information from the touchscreen self-filler questionnaire was linked to the health insurance information ‘privately insured’ and ‘name of the health insurance company’ (see **Table 1**) from the informed consent. Information from both data sources was compared and, if necessary, a correction was made in the self-reported variable. Validation was only possible for participants who provided information in both data sources (n=85,644; 41.7%). In implausible cases, the name of the health insurance company was used for validation. This procedure was used to derive a corrected variable for the self-reported health insurance status, which still has a high proportion of missing values due to the above-mentioned reasons.

The aim of our study was to develop and internally validate models to predict the health insurance status of participants in the NAKO baseline survey in order to replace missing values. In this respect, our research interest was focused on the question to which extent socio-demographic characteristics are suitable for predicting the health insurance status of participants in the NAKO for whom neither self-reports on health insurance status nor health insurance information from informed consent are available.

## Methods

### Database

During the baseline survey, 205,264 participants aged between 20 and 69 years were recruited between March 2014 and September 2019 in 18 study centres distributed throughout Germany. Sex- and age-stratified random samples (women and men with a share of 50% each; 10% each of 20-29 and 20-39 year-olds, 26.6% each of 40-49, 50-59 and 60-69 year-olds) were drawn from the general population via the regional population registers [1–4]. Further inclusion criteria were sufficient German language skills and the ability to give informed consent to participate in the study. CAPI’s, touchscreen self-filler questionnaires and physical ex-aminations were conducted. Certified and trained personnel as well as a common study protocol ensured standardised procedures applied by all study centres. The preliminary mean response rate was approximately 18% [2]. Further details on the study design and the study population can be found in [1–4, 18].

The present analysis was based on the data set generated from the NAKO baseline survey described above, where it should be noted that, with the exception of the variable on selfreported health insurance status, this is a non-quality assured data set. Nevertheless, initial plausibility checks indicate that the data quality is high. The data set also includes persons older than 69 years (n=4,401), since in some cases several years passed between sampling of participants and conduct of the baseline survey [27].

### Outcome variables

We developed two prediction models to estimate the probability that a participant is either member of a SHI (model 1) or PHI (model 2). Based on the operationalisation of the selfreported health insurance status (**Table 1**) we defined the outcome variable in model 1 as follows: 1=’statutorily insured’, 0=’not statutorily insured’. The category ’not statutorily insured’ includes participants who are privately, otherwise, or not health insured. In model 2, we used the following coding: 1=’privately insured’, 0=’not privately insured’. Participants who indicated having a statutory, other, or no health insurance were assigned to the category ‘not privately insured’.

### Predictor variables

We selected the predictors based on literature research. Due to the regulations for having access to PHI described above, there is a selection in PHI towards people with a higher average income and thus a higher socio-economic status by design. Various empirical studies have examined the distribution of socio-demographic differences between persons with SHI and PHI. These studies have shown that privately insured people have a higher socio-economic status in terms of income, education level and occupation compared to people with SHI. Besides, a comparatively higher proportion of women and elderly people are covered by SHI. The share of PHI-insured persons is higher in West Germany than in East Germany. In addition, there are differences in the family structures between the two groups of differently insured persons, since married persons are more likely to opt for SHI [7, 10–12, 15–17]. In summary, we identified eight potentially suitable predictors of health insurance status by the literature research: occupation, income, education, sex, age, employment status, residential area, and marital status. Other potentially relevant predictors of health insurance status, such as health status or migration background, were not considered because they were not included in the available data set.

The elicitation of socio-demographic characteristics in the NAKO was mainly based on the Federal Statistical Office’s *demographic standards* of 2010 [28]. Further information on the instruments used to measure socio-demographic characteristics in the NAKO and on their distribution at the half-time of the baseline assessment can be found in [18]. We included age as a continuous variable in the prediction models to avoid loss of information through classification. For descriptive purposes, we additionally classified age into 10-year groups analogously to the sampling strategy [2]. All other predictors were per se categorical variables. We divided the household income into five quantiles according to the recommendation of *demographic standards* [28]. The variable study centre served as a proxy for the residential area. Employment status was classified according to the International Labour Organisation (ILO) *Labour Force Concept* [29]. We combined the highest educational and vocational qualifications of the respondents based on the *Comparative Analysis of Social Mobility in Industrial Nations (CASMIN)* educational classification [28, 30, 31]. Additionally, we summarised the categories for the variables employment status and marital status further. The exact classifications of the respective variables are shown in **Table 2** to **Table 4**. Reporting of this study is based on the *Transparent Reporting of a multivariable prediction model for Individual Prognosis Or Diagnosis (TRIPOD)*-*G*uidelines [32, 33].

**Table 2.**
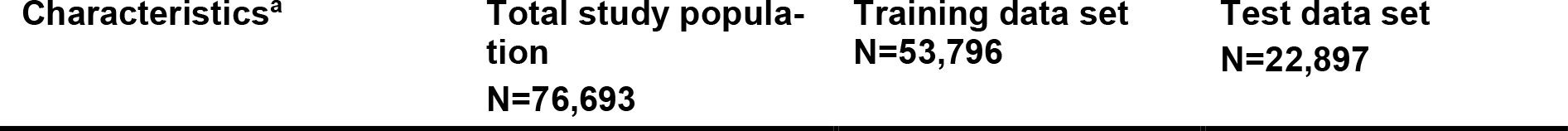
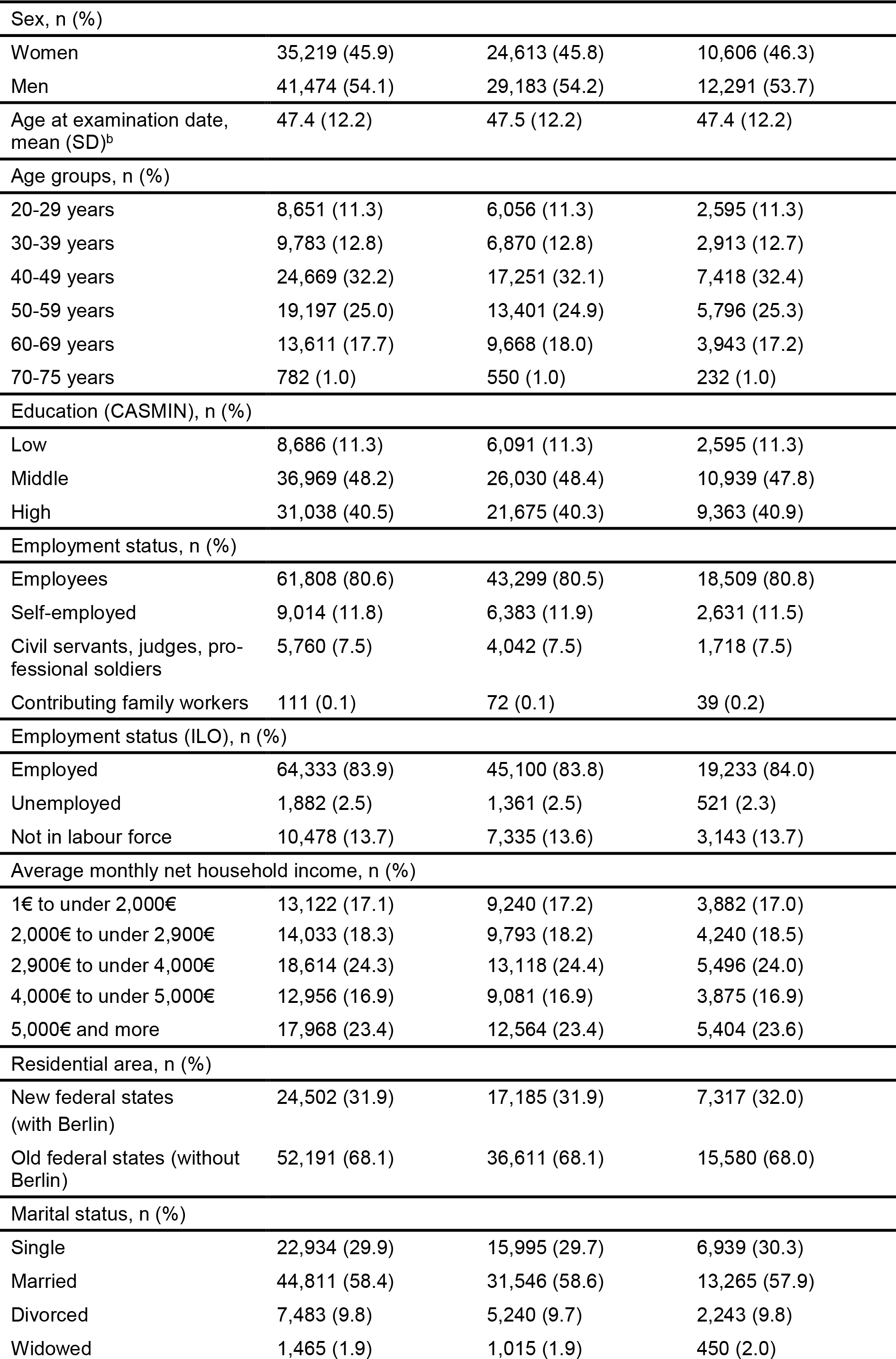
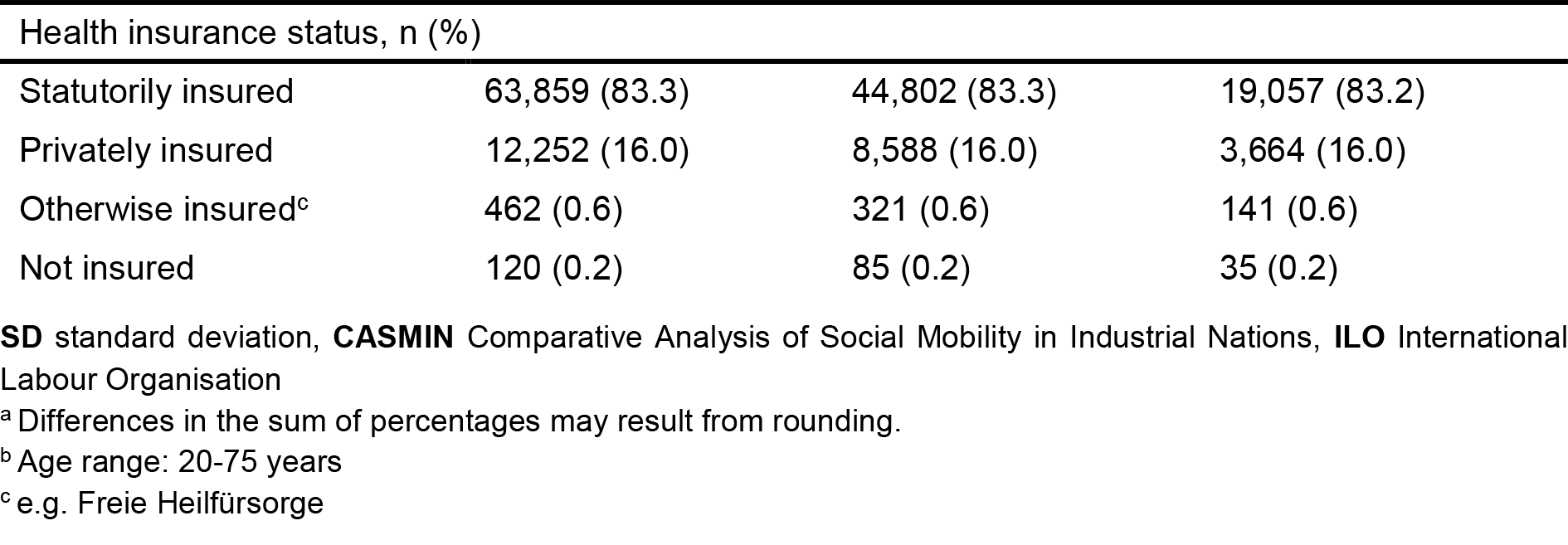
Description of the study population (German National Cohort) according to socio-demographic characteristics.

**Table 3.** Prediction model for the probability of membership in a SHI.

| Model estimates in the training data set<sup>a</sup> |  |  |
| --- | --- | --- |
| n | 53,796 |  |
| Number of SHI-insured persons | 44,802 |  |
| Variable | Beta Coefficient (SE) | p-value |
| Sex |  |  |
| Women | 0.874 (0.037) | <0.001 |
| Men | Ref. | Ref. |
| Age (per 1 year increase) | -0.031 (0.002) | <0.001 |
| Education (CASMIN) |  |  |
| Low | 0.320 (0.070) | <0.001 |
| Middle | Ref. | Ref. |
| High | -0.486 (0.036) | <0.001 |
| Employment status |  |  |
| Employees | Ref. | Ref. |
| Self-employed | -1.953 (0.036) | <0.001 |
| Civil servants, judges, professional soldiers | -5.655 (0.078) | <0.001 |
| Contributing family workers | -1.370 (0.356) | <0.001 |
| Employment Status (ILO) |  |  |
| Employed | Ref. | Ref. |
| Unemployed | 0.364 (0.161) | 0.024 |
| Not in labour force | -0.191 (0.059) | 0.001 |
| Average monthly net household income |  |  |
| 1€ to under 2,000€ | 1.038 (0.077) | <0.001 |
| 2,000€ to under 2,900€ | 0.513 (0.064) | <0.001 |
| 2,900€ to under 4,000€ | Ref. | Ref. |
| 4,000€ to under 5,000€ | -0.410 (0.055) | <0.001 |
| 5,000€ and more | -1.507 (0.047) | <0.001 |
| Residential area |  |  |
| New federal states (with Berlin) | 0,039 (0,037) | 0,293 |
| Old federal states (without Berlin) | Ref. | Ref. |
| **Marital status** |  |  |
| Single | -0,534 (0,045) | <0,001 |
| Married | Ref. | Ref. |
| Divorced | -0,414 (0,061) | <0,001 |
| Widowed | -0,474 (0,137) | 0,001 |
| Intercept | 4,681 (0,106) | <0,001 |
| **Assessment of the predictive performance in the test data set** |  |  |
| n | 22,897 |  |
| Number of SHI-insured persons | 19,057 |  |
| AUC (95%-CI) | 0.91 (0.91-0.92) |  |
| Calibration slope (95%-CI) | 0.97 (0.97-0.97) |  |

**SE** standard error, **CASMIN** Comparative Analysis of Social Mobility in Industrial Nations, **ILO** International Labour Organisation, **AUC** Area Under the Curve
<sup>a</sup> The predicted probability of a participant of being statutorily insured can be calculated as follows: $P(SHI=1) = 1/[1+\exp(-(4.681 + 0.874 \cdot \text{sex women} - 0.031 \cdot \text{age} + 0.320 \cdot \text{education low} - 0.486 \cdot \text{education high} - 1.953 \cdot \text{employment status self-employed} - 5.655 \cdot \text{employment status civil servants, judges, professional soldiers} - 1.370 \cdot \text{employment status contributing family workers} + 0.364 \cdot \text{employment status unemployed} - 0.191 \cdot \text{employment status not in labour force} + 1.038 \cdot \text{household income 1€ to under 2,000€} + 0.513 \cdot \text{household income 2,000€ up to under 2,900€} - 0.410 \cdot \text{household income 4,000€ up to under 5,000€} - 1.507 \cdot \text{household income 5,000€ and more} + 0.039 \cdot \text{residential area new federal states} - 0.534 \cdot \text{marital status single} - 0.414 \cdot \text{marital status divorced} - 0.474 \cdot \text{marital status widowed}))]$ . For categorical variables, a 1 is used if the predictor value is present and a 0 is used if it is absent.

**Table 4.** Prediction model for the probability of membership in a PHI.

|  |  |  |
| --- | --- | --- |
| **Model estimates in the training data set<sup>a</sup>** |  |  |
| n | 53,796 |  |
| Number of PHI-insured persons | 8,588 |  |
| **Variable** | **Beta Coefficient (SE)** | **p-value** |
| **Sex** |  |  |
| Women | -0.725 (0.035) | <0.001 |
| Men | Ref. | Ref. |
| Age (per 1 year increase) | 0.035 (0.002) | <0.001 |
| **Education (CASMIN)** |  |  |
| Low | -0.273 (0.069) | <0.001 |
| Middle | Ref. | Ref. |
| High | 0.599 (0.035) | <0.001 |
| **Employment status** |  |  |
| Employees | Ref. | Ref. |
| Self-employed | 1.974 (0.037) | <0.001 |
| Civil servants, judges, professional soldiers | 4.603 (0.057) | <0.001 |
| Contributing family workers | 1.193 (0.404) | 0.003 |
| **Employment Status (ILO)** |  |  |
| Employed | Ref. | Ref. |
| Unemployed | -0.535 (0.172) | 0.002 |
| Not in labour force | 0.246 (0.056) | <0.001 |
| Average monthly net household income |  |  |
| 1€ to under 2,000€ | -1.053 (0.076) | <0.001 |
| 2,000€ to under 2,900€ | -0.500 (0.061) | <0.001 |
| 2,900€ to under 4,000€ | Ref. | Ref. |
| 4,000€ to under 5,000€ | 0.367 (0.054) | <0.001 |
| 5,000€ and more | 1.471 (0.046) | <0.001 |
| Residential area |  |  |
| New federal states (with Berlin) | -0.112 (0.036) | 0.002 |
| Old federal states (without Berlin) | Ref. | Ref. |
| Marital status |  |  |
| Single | 0.524 (0.044) | <0.001 |
| Married | Ref. | Ref. |
| Divorced | 0.401 (0.059) | <0.001 |
| Widowed | 0.434 (0.132) | 0.001 |
| Intercept | -4.951 (0.104) | <0.001 |
| **Assessment of the predictive performance in the test data set** |  |  |
| n | 22,897 |  |
| Number of PHI-insured persons | 3,664 |  |
| AUC (95%-CI) | 0.91 (0.90-0.91) |  |
| Calibration slope (95%-CI) | 0.97 (0.97-0.97) |  |

**SE** standard error, **CASMIN** Comparative Analysis of Social Mobility in Industrial Nations, **ILO** International Labour Organisation, **AUC** Area Under the Curve
<sup>a</sup> The predicted probability of a participant of being privately insured can be calculated as follows: $P(\text{PHI}=1) = 1/[1 + \exp(-(-4.951 - 0.725 \cdot \text{sex women} + 0.035 \cdot \text{age} - 0.273 \cdot \text{education low} + 0.599 \cdot \text{education high} + 1.974 \cdot \text{employment status self-employed} + 4.603 \cdot \text{employment status civil servants, judges, professional soldiers} + 1.193 \cdot \text{employment status contributing family workers} - 0.535 \cdot \text{employment status unemployed} + 0.246 \cdot \text{employment status not in labour force} - 1.053 \cdot \text{household income 1€ to under 2,000€} - 0.500 \cdot \text{household income 2,000€ up to under 2,900€} + 0.367 \cdot \text{household income 4,000€ up to under 5,000€} + 1.471 \cdot \text{household income 5,000€ and more} - 0.112 \cdot \text{residential area new federal states} + 0.524 \cdot \text{marital status single} + 0.401 \cdot \text{marital status divorced} + 0.434 \cdot \text{marital status widowed}))]$ . For categorical variables, a 1 is used if the predictor value is present and a 0 is used if it is absent.

### Statistical analysis

We calculated absolute and relative frequencies for categorical variables and mean values and standard deviations for continuous variables to describe the study population. The analysis was conducted according to the approach proposed by Moons et al.: identification of predictors, regression analysis, assessment of predictive performance, and validation [34]. The predictive analysis consisted of two main steps: first, the development of the prediction models in the training data set, and, second, the internal validation in the test data set. We used a split-sample approach to avoid overfitting. The training data set comprised 70% and the test data set 30% of the data. Participants with missing data (4.4%) were deleted in both data sets (complete-case-analysis).

The first step of the statistical analysis was the development of the two prediction models. Using the full model approach, the predictor variables were included in prediction models. As already mentioned, we included all predictors by means of a priori knowledge. This procedure avoids overfitting, and a predictor selection bias [34].

The second step was to internally validate the prediction models. We calculated the predicted values by the two developed models in the test data set. The predictive performance was assessed considering discrimination and calibration. Discrimination describes the ability of a prediction model to distinguish between persons with and without outcome and was assessed using the Area Under the ROC Curve (AUC). The AUC takes values between 0.5 and 1 were an AUC of 0.5 indicates that the discriminative ability is not better than chance. An AUC of 1 corresponds to an ideal discrimination. In this study, the AUC represents the ability to distinguish between statutorily and not statutorily health insured persons and privately and not privately health insured persons. The Receiver Operating Characteristic (ROC) curve was used to visualise the discriminative ability. This is a graph showing the true positive rate (sensitivity) versus the false-positive rate (1 specificity).

Calibration means the agreement between the observed and predicted values. We assessed calibration with the calibration slope and graphically using the calibration plot [35]. In the calibration plot, we plotted the predicted probabilities against the observed values and added a line according to the Loess algorithm [36]. A diagonal 45° line was used for orientation and corresponds to an ideal calibration. We estimated the calibration slope with a logistic regression model by regressing the outcome on the logit of the predicted probability as the only predictor variable. A calibration slope of 1 indicates ideal calibration [35]. We calculated 95% confidence intervals for the performance parameters according to the TRIPOD-Guidelines [32]. The statistical analysis was done using IBM SPSS 26 ©.

## Results

### Selection of the study population

We excluded nine subjects due to implausible values in the age variable. A further 119,619 subjects were excluded, where only the health insurance information from the informed consent or only from self-reports was available, or no information on health insurance status was available in either data source. After excluding 8,943 subjects due to missing values in the outcome and predictor variables, the study population consisted of 76,693 persons. These were randomly assigned to a training data set (n=53,796) and a test data set (n=22,897) (**Fig. 2**).

**Fig. 2.**
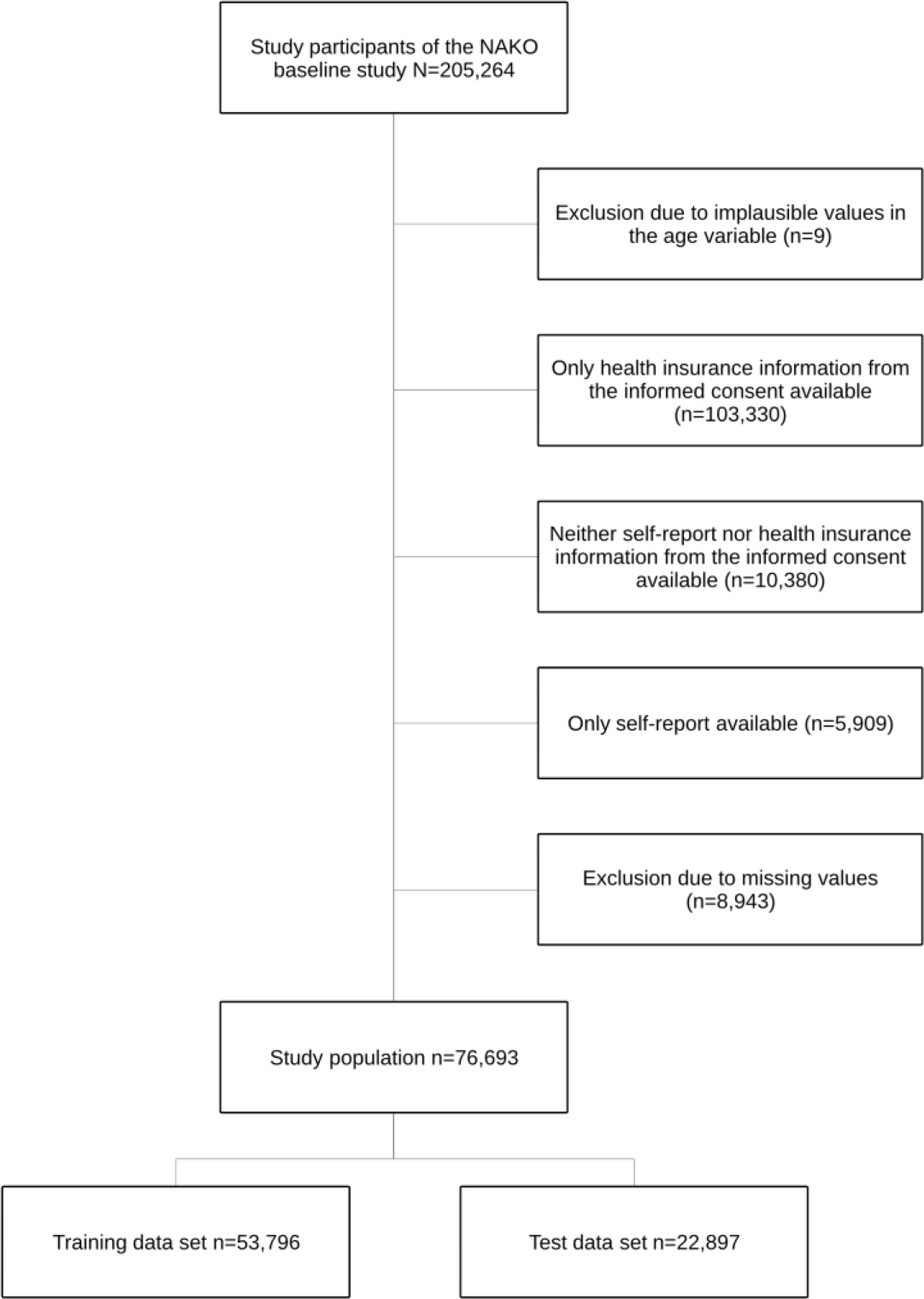
Selection of study population recruited 2019 - 2014 for the NAKO with 18 study centres

### Description of the study population according to socio-demographic characteristics

**Table 2** shows the socio-demographic characteristics of the total study population as well as the participants in the training and test data set (mean age 47 years). The proportion of men was higher than that of women (54% vs. 46%). In the training and in the test data set, 83.3% and 83.2% of the participants were statutorily health insured, and 16.0% were privately health insured. The proportions of otherwise insured and uninsured persons were less than 1% each.

### Prediction model for the probability of membership in a SHI

The prediction model for the probability of being insured by SHI and the performance of the model are shown in **Table 3**. We based the model on 53,796 participants, 44,802 of whom are insured in the SHI system. The most important predictors were employment status and household income. The residential area was left in the model despite a non-significant regression coefficient since other studies have shown regional differences between the two groups of differently insured persons.

The AUC of 0.91 (95%-CI: 0.91-0.92) indicated almost ideal discrimination between persons with SHI and non-SHI (**Table 3**). The ROC curve also showed the model’s good discriminative ability (**Fig. 3**). The calibration plot, which represents the agreement between observed and predicted values for membership in a SHI, showed an almost ideal calibration (**Fig. 4**). The calibration slope of 0.97 (95% CI: 0.97-0.97) did not show any overfitting problems (**Table 3**). Therefore, a correction of the regression coefficients was not necessary.

**Fig. 3.**
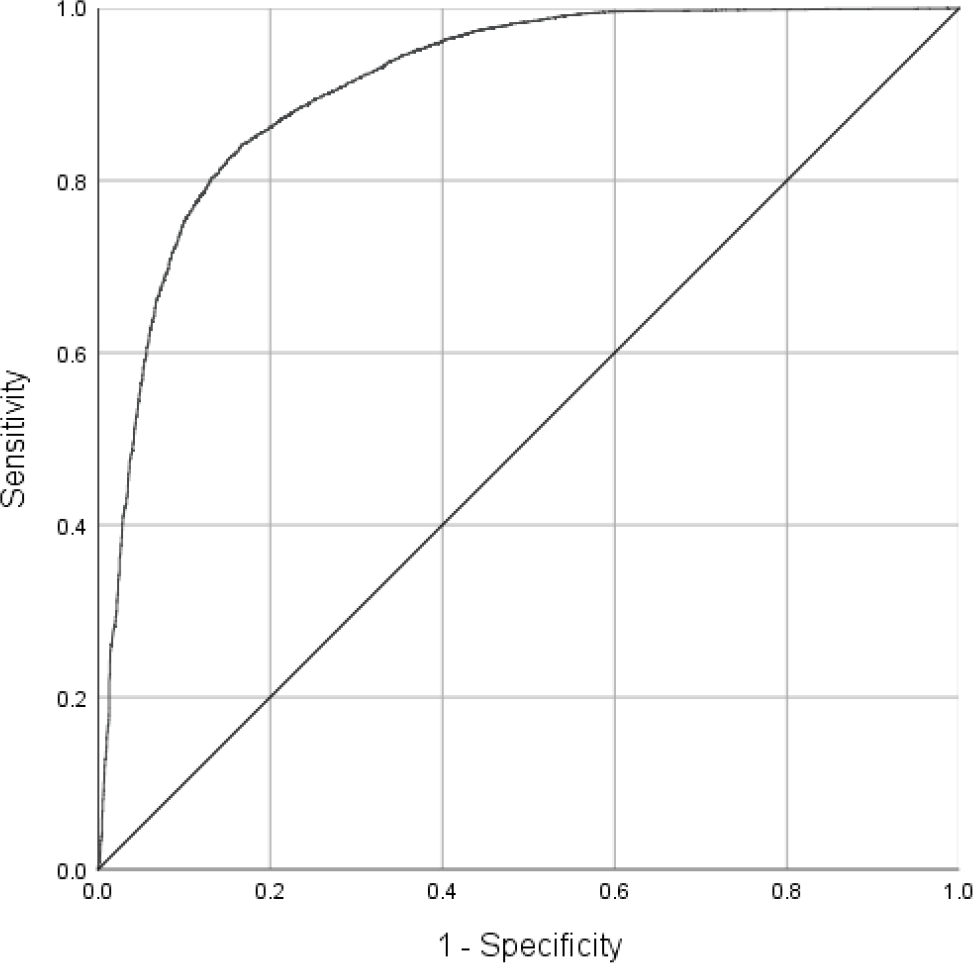
ROC curve for the prediction model for the probability of membership in a SHI

**Fig. 4.**
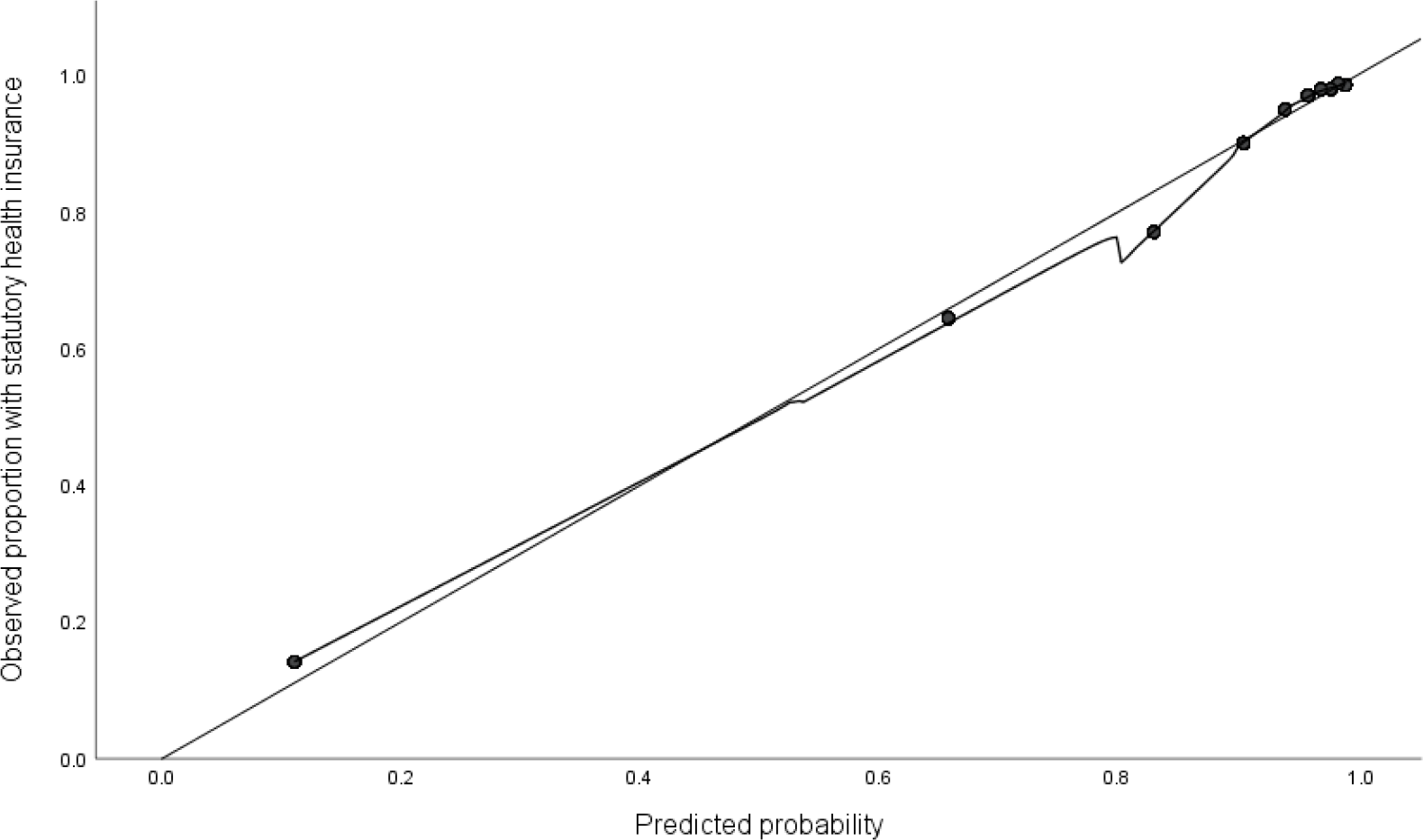
Calibration plot for the prediction model for the probability of membership in a SHI

### Prediction model for the probability of membership in a PHI

The prediction model for the probability of being insured by PHI and the performance of the model are shown in **Table 4**. We based the model on 53,796 participants, 8,588 of whom are insured in the PHI system. As in the first model, employment status and household income turned out to be important predictors (**Table 4**). According to the AUC of 0.91 (95% CI: 0.90- 0.91), this model had a very high discriminative ability, which was also shown in the ROC curve (**Fig. 5**). The calibration slope of 0.97 (0.97-0.97) and the calibration plot showed close to ideal calibration (**Fig. 6** & **Table 4**). The probabilities predicted by the model differed only slightly from the observed values.

**Fig. 5.**
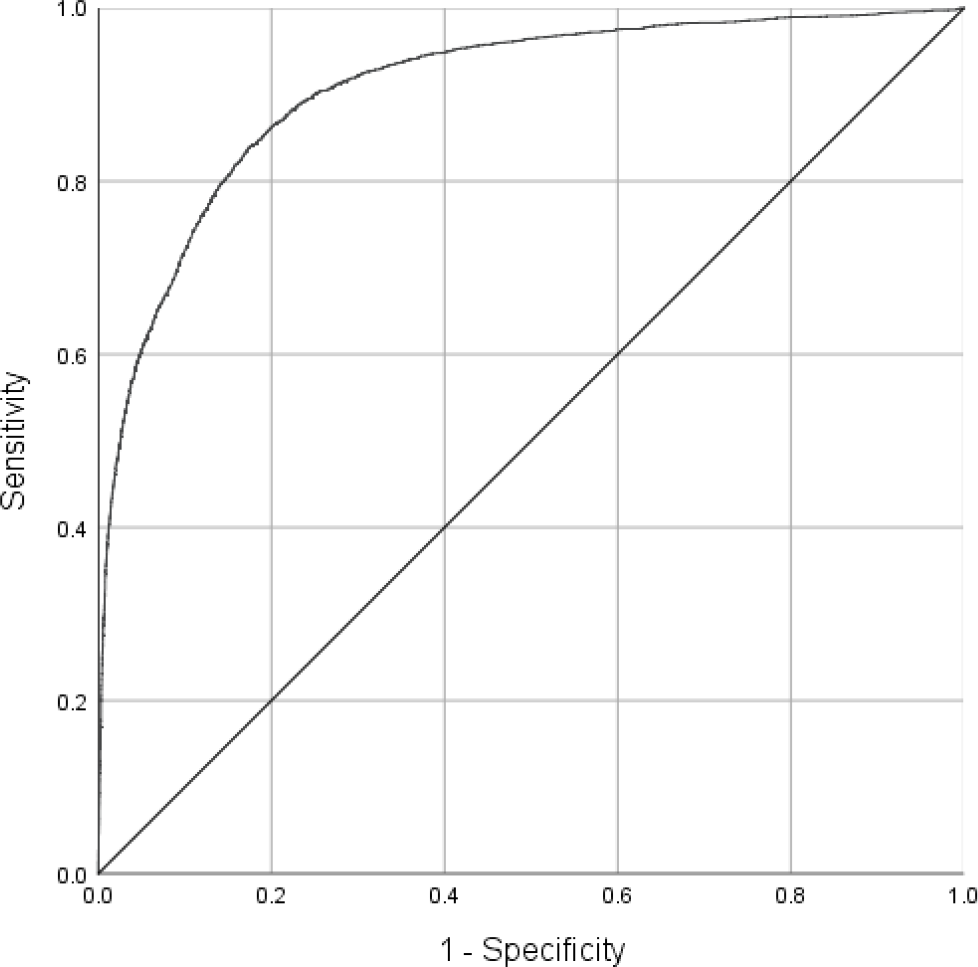
ROC curve for the prediction model for the probability of membership in a PHI

**Fig. 6.**
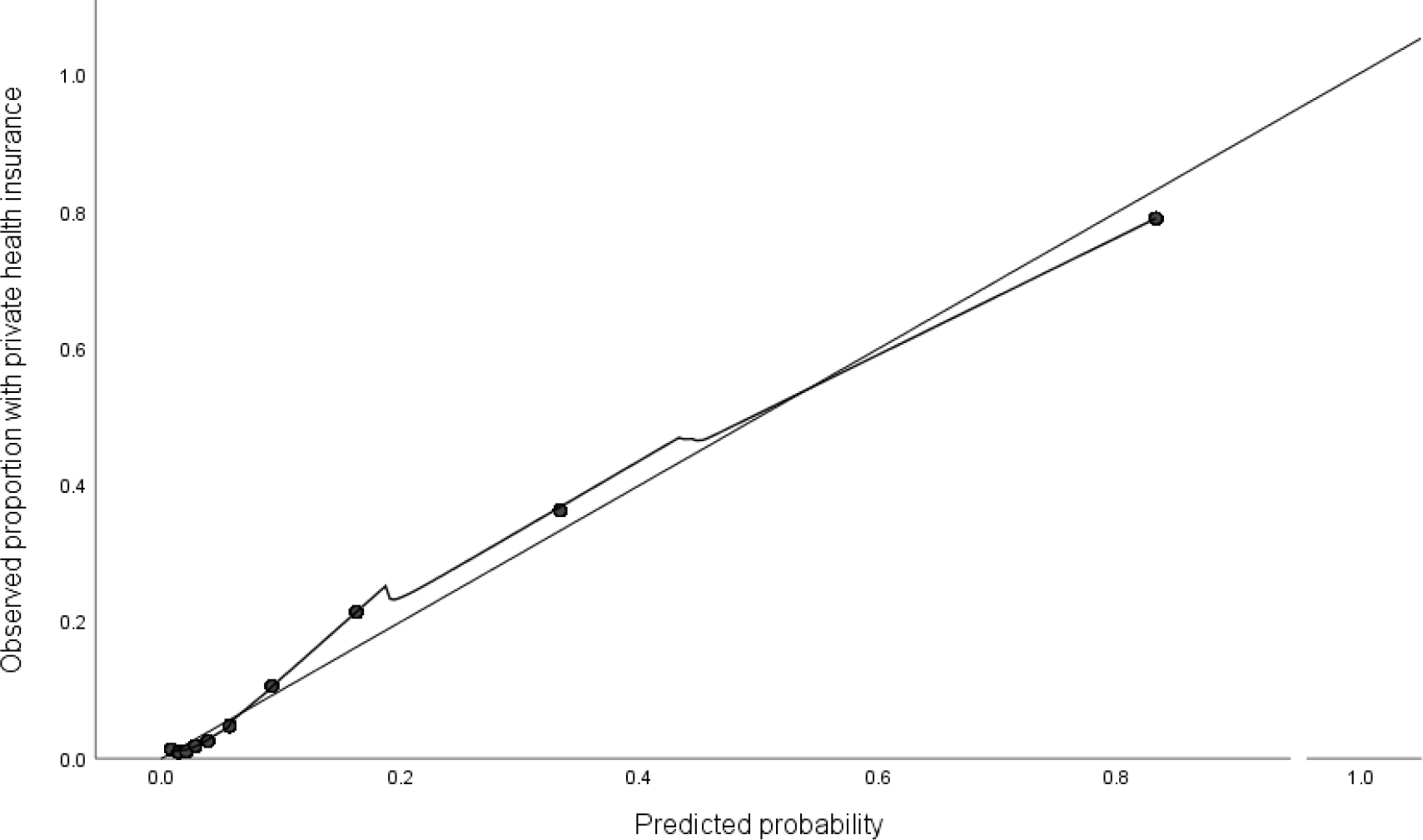
Calibration plot for the prediction model for the probability of membership in a PHI

## Discussion and Conclusions

### Key findings

The present study aimed at answering the question to which extent selected socio-demographic characteristics are suitable for predicting the health insurance status of participants in the NAKO baseline survey for whom neither self-reports on health insurance status nor health insurance information from informed consent are available. For this purpose, we developed and internally validated two prediction models. We investigated the performance of the models with respect to their discrimination and calibration ability to assess whether the predicted values can be used as reliable replacement of the missing values in the variable on self-reported health insurance status.

Information on the health insurance status is available from participants who have agreed to provide claims data via their health insurance. In addition, the self-reported health insurance status has been collected during the baseline survey since 2017. The variable on self-reported health insurance status has a high proportion of missing values due to the subsequent inclusion of the question in the touchscreen self-filler questionnaire during its revision. For 5.1% of the participants, neither of the two data sources contains information on health insurance status.

The literature review identified occupation, income, education, sex, age, employment status, residential area, and marital status as potentially suitable predictors of health insurance status [7, 10–12, 15–17]. Based on this information, we developed and internally validated two prediction models. Model 1 estimated the probability of a person being insured by SHI and model 2 estimated the probability of a person being insured by PHI. The internal validation showed extraordinarily good performance of the developed prediction models. Based on performance parameters and via graphical representations both models turned out to show an almost ideal discrimination and calibration. The models distinguished very well between persons with and without the respective outcome (SHI and PHI). The calibration plots showed that the probabilities predicted by the models differ only slightly from the observed values. In model 1, the observed values were slightly lower than the predicted probabilities. Model 2 showed the opposite picture. External validation is necessary for further assessment of their calibration, since here, for example, the calibration-in-the-large can also be considered additionally [35].

The results of the internal validation clearly show that the socio-demographic characteristics included in the models prove to be suitable predictors for the health insurance status of the participants in the NAKO baseline survey. In particular, employment status and household income are important to predict the health insurance status of NAKO participants. This finding is very plausible considering the regulations for having access to PHI. PHI only insures persons with a gross income above the opt-out threshold or specific professional groups such as civil servants or self-employed [8]. It should be noted that in the present study, the monthly net household income was included in the analyses, as the NAKO does not collect respondent income.

## Strengths and limitations

The strengths of our analysis included the large study population drawn from random samples of regional population registers and the high number of outcomes, which significantly influence the robustness of statistical results in predictive analyses. In addition, large samples reduce the probability of an overly optimistic estimate of the predictive performance [32, 35]. A further strength was the standardised collection of the predictor variables. On the one hand, this ensured high data quality with regard to the socio-demographic characteristics in the NAKO [18]. On the other hand, the orientation towards the *demographic standards* in the collection of the characteristics enables a certain reproducibility. The models developed can be applied to data sets or studies in which the socio-demography of the participants is acquired in the same way. Using the equations given in **Table 3** and **Table 4**, the predicted probability of membership in SHI or PHI can be calculated. Besides, the prediction models were developed and internally validated considering current recommendations and guidelines.

The present analysis also has some limitations. First, the lack of external validation of the prediction models means that the results may not be generalised to other research settings. Second, other potentially relevant predictors of health insurance status, such as health status or migration background, were not considered because they were not included in the available data set. Another limitation was the dichotomisation of the outcome variables. The development of a model for the prediction of all possible health insurance statuses could have been realized using multinomial logistic regression. This would be of interest for an optimisation or completion of the variable on the self-reported health insurance status. In the present study, only the outcomes SHI and PHI were considered, since the literature on predictive modelling mainly refers to binary endpoints. Additionally, in the context of e.g. health services research, the focus lies on the distinction between those with SHI and PHI.

### Implications and recommendations for future research

Our findings show that socio-demographic characteristics are suitable predictors for the health insurance status of the participants in the NAKO baseline survey. The predicted values can be used as reliable replacement of the missing values in the variable on self-reported health insurance status. However, before the models are used, e.g. for the preparation and processing of data from other studies, an external validation in population-based studies is recommended.

Future studies could investigate to which extent replacing the missing values in the variable on the self-reported health insurance status with the developed prediction models differs from multiple imputation and which procedure yields better results.

## Data Availability

The data that support the findings of this study are available from the German National Cohort but restrictions apply to the availability of these data, which were used under license for the current study, and so are not publicly available.

## List of abbreviations

AUC: Area Under the Curve; CAPI: Computer-assisted personal interview; CASMIN: Comparative Analysis of Social Mobility in Industrial Nations; EPV: Events per variable; ILO: International Labour Organization; NAKO: German National Cohort; PHI: Private health insurance; ROC curve: Receiver Operating Characteristic-Curve; SD: Standard deviation; SE: Standard error; SHI: Statutory health insurance; TRIPOD: Transparent Reporting of a multivariable prediction model for Individual Prognosis Or Diagnosis

## Declarations

### Ethics approval and consent to participate

The study protocol of the NAKO was approved by the ethics committee of the Bavarian State Medical Association (13023 and 13031) and by the locally responsible ethics committees of the institutions of the 18 study centres. All the described investigations were conducted in compliance with national law and in accordance with the declaration of Helsinki (in the latest revised version). All participants have been fully informed and have given their written informed consent to participate in the study.

### Consent for publication

Not applicable

### Competing interests

The authors declare that they have no competing interests.

### Funding

This project was conducted with data from the German National Cohort (NAKO) (www.nako.de). The NAKO is funded by the Federal Ministry of Education and Research (BMBF) [project funding reference numbers: 01ER1301A/B/C and 01ER1511D], federal states and the Helmholtz Association with additional financial support by the participating universities and the institutes of the Leibniz Association. We thank all participants who took part in the German National Cohort and the staff in this research program.

### Authors’ contributions

ES, CS and IH conceptualised the study. HB1, HB2, ADM, WH, KHJ, NK, VK, TK, BK, ML, CMF, KM, RM, TN, IP, SS, BS, BW, SW, RW, ES and CS were responsible for data curation. CS did the data cleaning and created the final dataset. IH conducted the statistical analysis and wrote the original draft of the manuscript. CS and ES supervised IH. HB2, VK, TK, BK, CMF, RM, TN, IP, SS, CS and ES substantively revised the original draft of the manuscript. All authors read and approved the final manuscript.

## Acknowledgements

This article presents a translated part of the first author’s master’s thesis as a slightly modified version, initially written in the German language and bearing the following title: Entwicklung und interne Validierung von Prognosemodellen zur Vorhersage des Krankenversicherungssta- tus von Teilnehmer*innen der Basiserhebung der NAKO Gesundheitsstudie. Magdeburg, Berlin School of Public Health, 2020.

We would like to thank Christine Wallisch for her methodical consulting. Further, we are grateful to Ulrike Nimptsch for advice in the planning of the study.

## Additional Information

Transparent Reporting of a multivariable prediction model for Individual Prognosis Or Diagnosis (TRIPOD)-Guidelines [37]

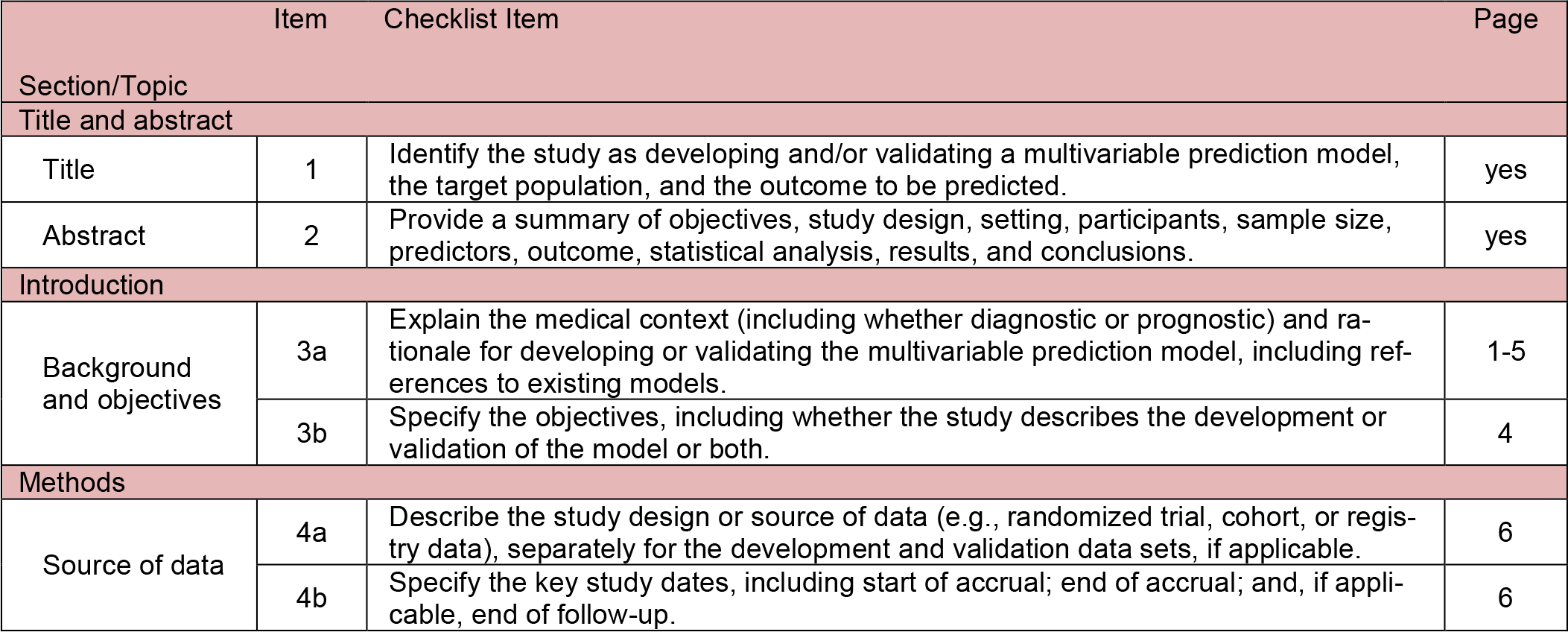
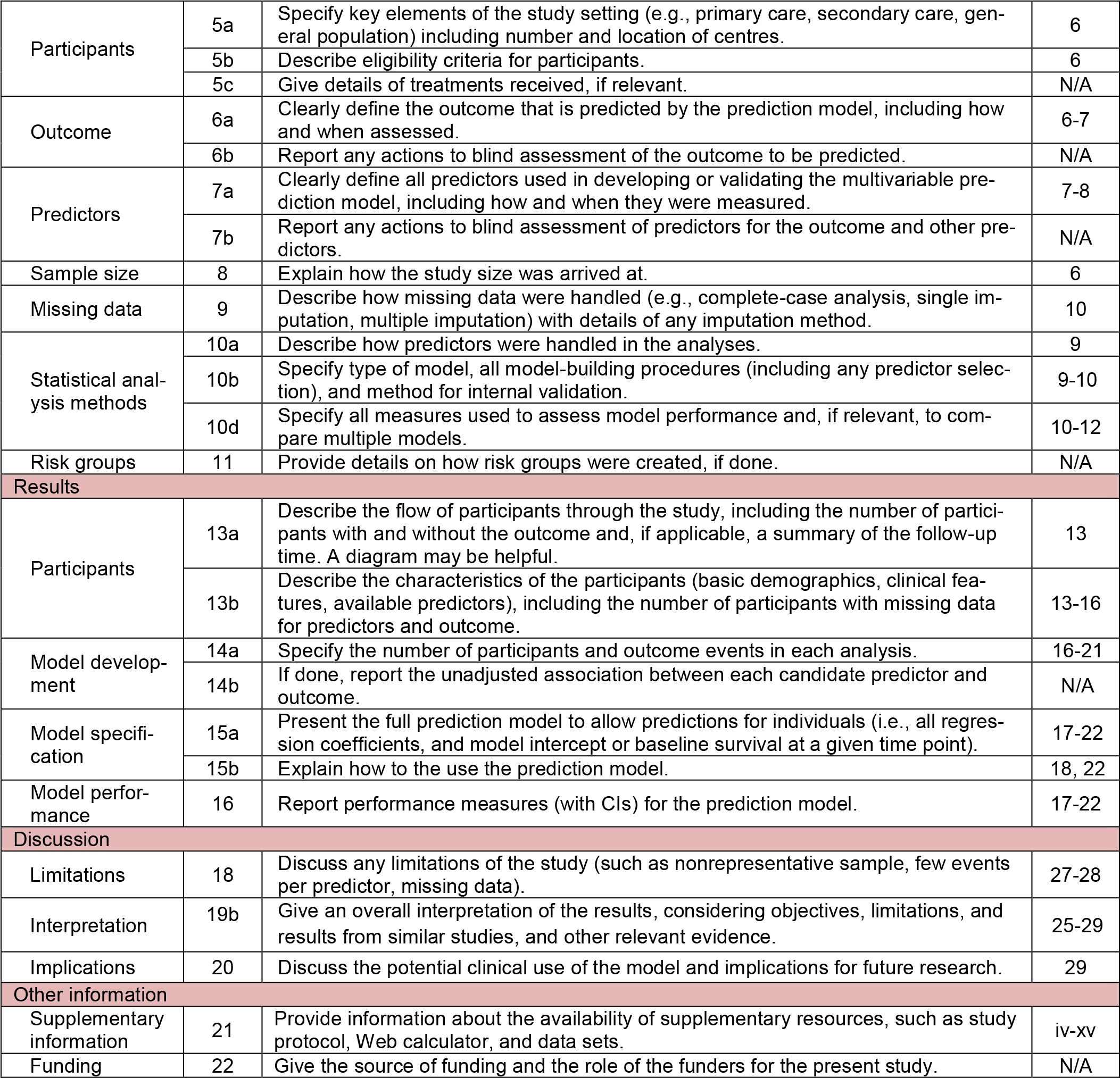

## Notes

### Competing Interest Statement

The authors have declared no competing interest.

